# Quantifying the genetic contributions in unexplained kidney failure reveals *APOL1-HLA* interaction

**DOI:** 10.1101/2024.10.10.24315080

**Authors:** Omid Sadeghi-Alavijeh, Melanie MY Chan, Konstantinos Tzoumkas, Gabriel T. Doctor, Daniel P Gale

**Affiliations:** Centre for Genetics and Genomics, UCL Department of Renal Medicine, UCL Medical School, Rowland Hill Street, London NW3 2PF; Medical Research Council Laboratory of Medical Sciences, Hammersmith Campus, Du Cane Road, Imperial College London, W12 0NN

**Author notes:** Corresponding author: Daniel P. Gale, Centre for Genetics and Genomics, UCL Department of Renal Medicine, UCL Medical School, Rowland Hill Street, London NW3 2PF. These authors contributed equally (Omid Sadeghi-Alavijeh, Melanie MY Chan).

**Keywords:** whole-genome sequencing, kidney failure, *APOL1*, genetics

## Abstract

**Background:** Unexplained kidney failure (uKF) affects 15% of individuals requiring kidney replacement therapy. Absence of a diagnosis creates uncertainty around recurrence after transplantation, familial risk, and participation in therapeutic trials. Whole genome sequencing (WGS) was used to identify genetic variants contributing to uKF.

**Methods:** 218 patients who presented with uKF < 50 years old were recruited to the UK’s 100,000 Genomes Project. Candidate variants in 183 genes were reviewed for pathogenicity by a multidisciplinary team. Gene-based association testing, structural variant analyses, and assessment of high-risk *APOL1* genotypes were performed. Polygenic risk scores (PRS) were calculated for chronic kidney disease (CKD), and various glomerulonephritides. HLA associations in those with *APOL1* high-risk genotype were also investigated.

**Results:** A positive genetic diagnosis was made in 17% (38/218) of patients. The median age of uKF onset was 36 years. Fewer genetic diagnoses were found in those aged ≥ 36 years compared to younger individuals, both with (11% vs. 35%, P=0.03) and without (5% vs. 19%, P=0.05) a family history. Three patients ≥ 36 years without a family history had pathogenic variants in type IV collagen genes. High-risk APOL1 genotypes were enriched in patients with recent African ancestry (52% vs 8.4%, P=5.97×10^−8^). Dividing the uKF cohort by subsequent identification of monogenic diagnosis, High-risk APOL1 genotype, or neither, we found that the SSNS PRS was higher in those with High-risk APOL1 (P=0.048), driven by differences at *HLA-DQB1*\**03:19* (P=0.001).

**Conclusions:** These findings estimate the likelihood of a genetic diagnosis using WGS in uKF patients, showing fewer diagnoses in older patients without a family history. *APOL1* contributes significantly to uKF in those with recent African ancestry, potentially interacting with *HLA-DQB1*. The lack of PRS signal for CKD suggests distinct biology between uKF and more common causes of CKD.

## Introduction

Chronic kidney disease (CKD) is often asymptomatic until damage to the kidneys is advanced. Consequently, patients (especially adults) frequently present with non-specific glomerulosclerosis and tubulointerstitial fibrosis on kidney biopsy which is uninformative as to the underlying cause of their disease. As a result, around 15-20% of adults who require chronic dialysis or kidney transplantation (kidney replacement therapy, KRT) have kidney failure that is unexplained (uKF)^1^. Even in patients with a diagnosis, rare diseases make a contribution to KF that is disproportionate to their frequency among patients with CKD when compared with more common causes of CKD such as diabetes^2^. The different contributions of common causes of CKD compared with rare disorders to KF among those in whom a diagnosis has not been made has not been established.

Over 450 genes have been identified that cause monogenic kidney disease with recent technological advances making gene panels, whole exome, or whole genome sequencing, available to large numbers of patients^3^. These technologies have shown that monogenic causes make a substantial contribution to the overall burden of kidney disease, identifying a molecular diagnosis in 9.3% of over 3300 patients with kidney disease/failure including 17% of those with nephropathy of unknown origin^4^. Exome and genome sequencing carried out in smaller cohorts of uKF report a diagnostic yield between 12-56% depending on the population studied ^5–9^.

Identifying the molecular cause of kidney disease in a patient can confirm the diagnosis, inform prognosis, enable predictive testing of family members, and facilitate transplant and reproductive decisions. Efforts to broaden availability of genomic testing are underway in many countries across the world. However, interpretation of genetic variants in known monogenic kidney disease genes can be challenging because of the frequent occurrence of rare (non – disease-causing) variants in individuals and so a high level of certainty is needed that the variant identified is responsible for a patient’s disease in order for it to be reported back to them. Variant interpretation is therefore a significant obstacle to delivery of medically actionable information from genetic tests. The overall likelihood that a medical test result is interpreted correctly is heavily dependent on the prior probability of that outcome, so information that allows prior probability to be estimated accurately is enormously valuable, assisting both the interpretation of candidate variants and policy decisions on which patient groups (as defined by clinical features) are most likely to benefit from genetic testing.

Here we use clinically accredited whole genome sequencing data from individuals with uKF recruited to the UK’s 100,000 Genomes Project to examine which clinical features are associated with an underlying monogenic disorder. In addition, we combine rare variant studies with analyses of the burden of known common genetic risk factors for different types of kidney disease (polygenic risk scores) to gain insight into the contribution of non-Mendelian diseases to uKF. This reveals evidence of a potential HLA driven autoimmune podocytopathy in individuals of recent African ancestry with uKF.

## Methods

### The 100,000 Genomes Project

The Genomics England dataset (version 10) includes whole-genome sequencing (WGS) data, clinical phenotypes encoded using Human Phenotype Ontology (HPO) codes, and NHS hospital records for 89,139 individuals recruited with cancer, rare disease, and their unaffected relatives. Ethical approval for the 100KGP was granted by the Research Ethics Committee for East of England – Cambridge South (REC Ref: 14/EE/1112).

### Unexplained kidney failure in young people

Unrelated probands recruited to the ‘Unexplained kidney failure in young people’ cohort in the 100,000 Genomes Project (100KGP) were identified. These are individuals who required KRT before the age of 50 years in the absence of an identified cause for their KF. Prior to recruitment, any patients with a personal or family history of gout before the age of 30 in the absence of CKD stage 3, 4 or 5, or diabetes, were expected to be tested for *UMOD* and *HNF1B* variants, respectively.

The exclusion criteria were: likely or proven diabetic nephropathy; likely or proven renovascular disease; identified glomerular disorder on kidney biopsy (other than glomerulocystic disease, ischaemic changes or secondary glomerulosclerosis); evidence of autoimmune, infectious, malignant, metabolic or other systemic disorder likely to be responsible for kidney disease; renal sarcoidosis or tuberculosis; paraproteinaemia (unless kidney biopsy shows no evidence of renal monoclonal deposition); exposure to nephrotoxin suspected of causing renal dysfunction; obstructive uropathy; significant proteinuria (>1g/day; uPCR >100) at presentation unless presentation was with kidney failure; identified tubular/electrolyte/acid base disorder; >5 kidney cysts (not attributable to acquired cystic disease of CKD); nephrolithiasis; structural kidney and urinary tract malformations.

DNA preparation and extraction, WGS, alignment and variant calling is described in detail in Supplementary Methods 1.

### Identification of pathogenic and likely pathogenic variants

Candidate variants identified within an expert-curated virtual gene panel of 183 (see supplementary table 1 for the full gene list) known kidney disease genes (https://panelapp.genomicsengland.co.uk/panels/678/) were assessed against American College of Medical Genetics and Genomics (ACMG) criteria to identify pathogenic or likely pathogenic variants^10^. These well-defined criteria integrate variant information including population frequency, computational predictions of deleteriousness, functional domain localisation, putative mechanism of disease and previous associations with phenotypes in reputable databases.

For the majority of patients, variants were discussed in a multidisciplinary team including clinical and molecular geneticists and the recruiting clinician. Outcomes were recorded as “solved,” “partially solved” or “not solved” in an *Exit Questionnaire* returned to Genomics England. This dataset was queried for patients from the uKF cohort and those taken as “solved” with respect to their kidney disease were placed into the solved category.

For ‘unsolved’ patients we analysed the output from the virtual gene panel applied to their WGS data looking for high impact variants (e.g. likely loss-of function) and de novo moderate impact variants (e.g. missense).

### Downstream bioinformatic analysis of the unsolved cohort

In the remaining unsolved uKF patients we attempted to ascertain whether there were missed genetic drivers by creating an uKF cohort ancestrally matched to a control cohort to conduct various collapsing analyses on. This cohort of 187 cases and 26,373 controls then underwent rare variant collapsing analysis using SAIGE-GENE, collapsing variants across a number of “masks” or filters. The masks used for this analysis were a rare, damaging missense mask (“missense+”), a high confidence loss of function mask (“LoF”), an intronic mask (“intronic”), a splice site mask, a 3-prime untranslated region mask (3’-UTR) and a 5-prime UTR mask (5’-UTR). We then applied an exome-wide structural variant analysis pipeline, using calls made by MANTA^11^ and CANVAS^12^ to look for structural and copy number variants that may account for uKF cases. Finally, we conducted polygenic risk score, and HLA analysis detailed below.

Full details of the cohort creation and collapsing analyses can be found in supplementary methods 3-5.

### *APOL1* and HLA analysis

Using genetically predicted ancestries calculated by the 100KGP, we divided the cases into those predicted with greater than 90% confidence to be of African ancestry (n=27) and those not (n=191) and created a subset of the controls who were of African ancestry without uKF (n=608). The *APOL1* G1 (S342G and I384M) and G2 (del388N389Y) renal risk variants were bioinformatically ascertained from WGS data for the cases and controls. Patients with a G2 allele who had the protective p.N264K missense variant ^13^, were reassigned as non-high risk *APOL1* genotype. The burden of high risk *APOL1* genotypes (G1/G1, G1/G2 and G2/G2) were compared across cohorts (case vs control, *APOL1* cases versus African ancestry controls with and without high risk *APOL1*) using a one-sided Fisher’s exact test and this variable (the presence or absence of a high-risk genotype) was used as part of the logistic regression model alongside various polygenic risk scores, the first ten principal components, age and sex.

The HLA types in cases and controls with high-risk *APOL1* genotype were then joint-analysed across the 100KGP and UK Biobank (UKBB) using a logistic regression model using HLA-type and five principal components, a chi-squared test was used to attribute for significance using a Bonferroni corrected P-value of 0.007 (7 HLA classes analysed, α=0.05). A full description can be found in Supplementary Methods 6.

### Polygenic risk scoring

Four polygenic risk scores (PRS) were applied to the unsolved cases and controls. A multi-ancestry IgA nephropathy score encompassing 77 variants^14^; a European membranous nephropathy score encompassing 12 variants^15^; a European steroid sensitive nephrotic syndrome (SSNS) score encompassing 5 variants^16^ and a multi-ancestry CKD score encompassing 471,316 variants^17^. The scores were lifted over from genome build 37 to 38 using the UCSC liftover tool^18^ and applied to the cohorts using the “score” command in PLINK2^19^. PRS scores were standardized to controls using Z-score scaling.

The uKF cases were divided into those with high-risk *APOL1* genotypes, those with a monogenic diagnosis delivered by the clinical arm of 100KGP and those patients who were unsolved. To test the significance between the PRS of cases and controls we applied an ANOVA test followed by a Tukey’s HSD test to differentiate the source of statistical significance using base R functions. All plotting was performed with ggplot2 in R^20^.

A logistic regression model with each PRS and the presence or absence of the high-risk *APOL1* genotype, sex, age and the first ten principal components was fitted to the unsolved uKF and control cohort in R and plotted in ggplot2.

### Statistics

Baseline characteristics are expressed as frequencies (n, %) and medians (IQR), as dictated by data type. A two-sided Fisher’s exact test was used to compare clinical characteristics of those with and without a genetic diagnosis. A Wilcoxon-Mann-Whitney test was used to compare age of KF between groups. Multivariable logistic regression was performed to identify clinical characteristics associated with a positive genetic result and the differences between PRS scores in cases and controls. All statistical analysis was performed in R (version 3.6.2). Two-sided *p* values < 0.05 were considered statistically significant.

## Results

We analysed WGS data from 218 probands with uKF: 62% were male, 40% had an affected first-degree relative, 53% had extra-renal manifestations, and 6% had self-reported parental consanguinity. Data on age at KF was available for 190/218 (87%) individuals with the median age at KF calculated as 36 years (IQR 16). 38/218 (17%) had a pathogenic or likely pathogenic variant affecting a known kidney disease gene, with 31/38 (82%) solved cases having the age at KF available (Figure 1, Supplementary Table 2).

**Figure 1.**
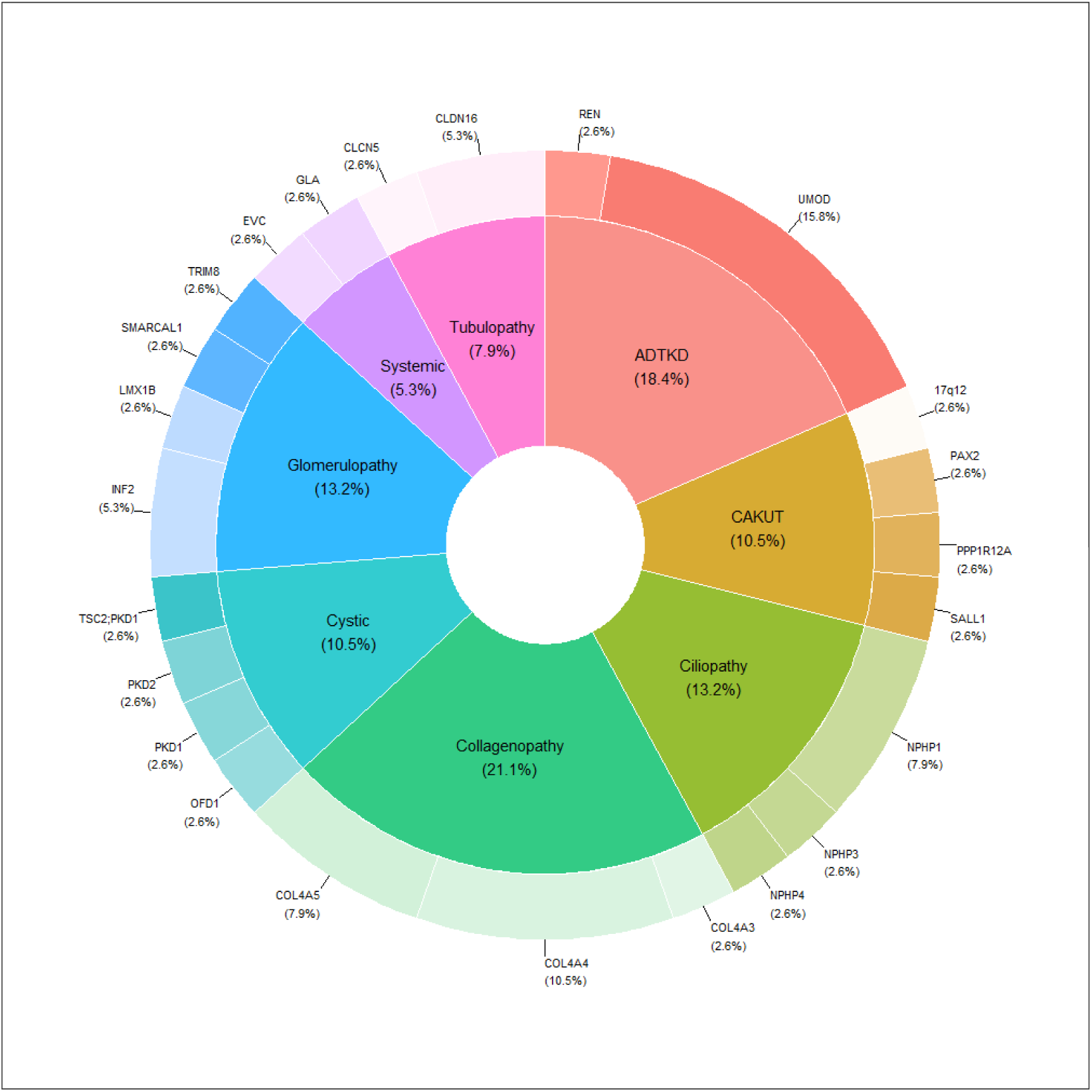
Breakdown of solved cases by gene category. Breakdown of the solved uKF cases by gene category. Percentages in the pie chart of the 38 solved cases, percentages within the doughnut represent percentages within their respective category. ADTKD, autosomal dominant tubulointerstitial kidney disease; CAKUT, congenital anomalies of the kidneys and urinary tract.

Pathogenic CNVs were seen in 2/218 (0.9%) individuals: A homozygous 110kb whole gene deletion of *NPHP1* was detected in a male with microscopic haematuria, proteinuria and gout who developed KF in their 30s (30-35). In addition, a 1.9Mb 17q12 duplication (encompassing *HNF1B*) was detected in a female with small kidneys who developed KF in their 30s (30-35). The duplication segregated with her affected mother.

### Clinical predictors of a positive genetic diagnosis

We next investigated whether there were any differences between patients with and without a molecular diagnosis to ascertain who might benefit most from genetic testing (Table 1). Clinical and demographic features were similar between the two groups and although median age at kidney failure was lower in those with a genetic diagnosis, this was not statistically significant (Table 1). Multivariable logistic regression did not identify any significant predictors of a positive genetic diagnosis in this relatively small cohort (Supplementary Table 3 and Supplementary Figure 2) and there was no association seen with specific extra-renal HPO terms (eye, ear, autoimmune, haematological, diabetes, endocrine, neurological, dermatological, gout, congenital malformations).

**Table 1.** Comparison of clinical and demographic features in patients with (solved) and without (unsolved) a genetic diagnosis. IQR, interquartile range; KF, kidney failure. P values calculated using a Mann Whitney Wilcoxon test (age at KF) or two-sided Fisher’s exact test.

|  | **Solved (n=38)** | **Unsolved (n=180)** | **P** |
| --- | --- | --- | --- |
| **Median age at KF (IQR)** | 31 (12) | 37 (16) | 0.11 |
| **Male sex (%)** | 24 (63%) | 111 (62%) | 1 |
| **Family history (%)** | 18 (47%) | 69 (38%) | 0.36 |
| **Extra-renal features (%)** | 25 (66%) | 90 (50%) | 0.11 |
| **Consanguinity (%)** | 3 (8%) | 9 (5%) | 0.44 |

The diagnostic yield in those with and without a family history was compared stratifying by age at KF (Table 2). In those with a family history, significantly fewer genetic diagnoses were made in those ≥ 36 years compared to those < 36 years (35% vs 11%, P=0.03) with the diagnostic yield in individuals ≥ 36 years without a family history just 5% (Figure 2).

**Table 2.** Diagnostic yield stratified by age of kidney failure and family history. KF, kidney failure. P values calculated using a two-sided Fisher’s exact test.

| **Family History** | **Age at KF < 36 years** | **Age at KF $\geq 36$ years** | **P** |
| --- | --- | --- | --- |
| **Yes** | 9/26 (35%) | 5/45 (11%) | 0.03 |
| **No** | 12/64 (19%) | 3/55 (5%) | 0.05 |
| **P** | 0.17 | 0.46 |  |

**Figure 2.**
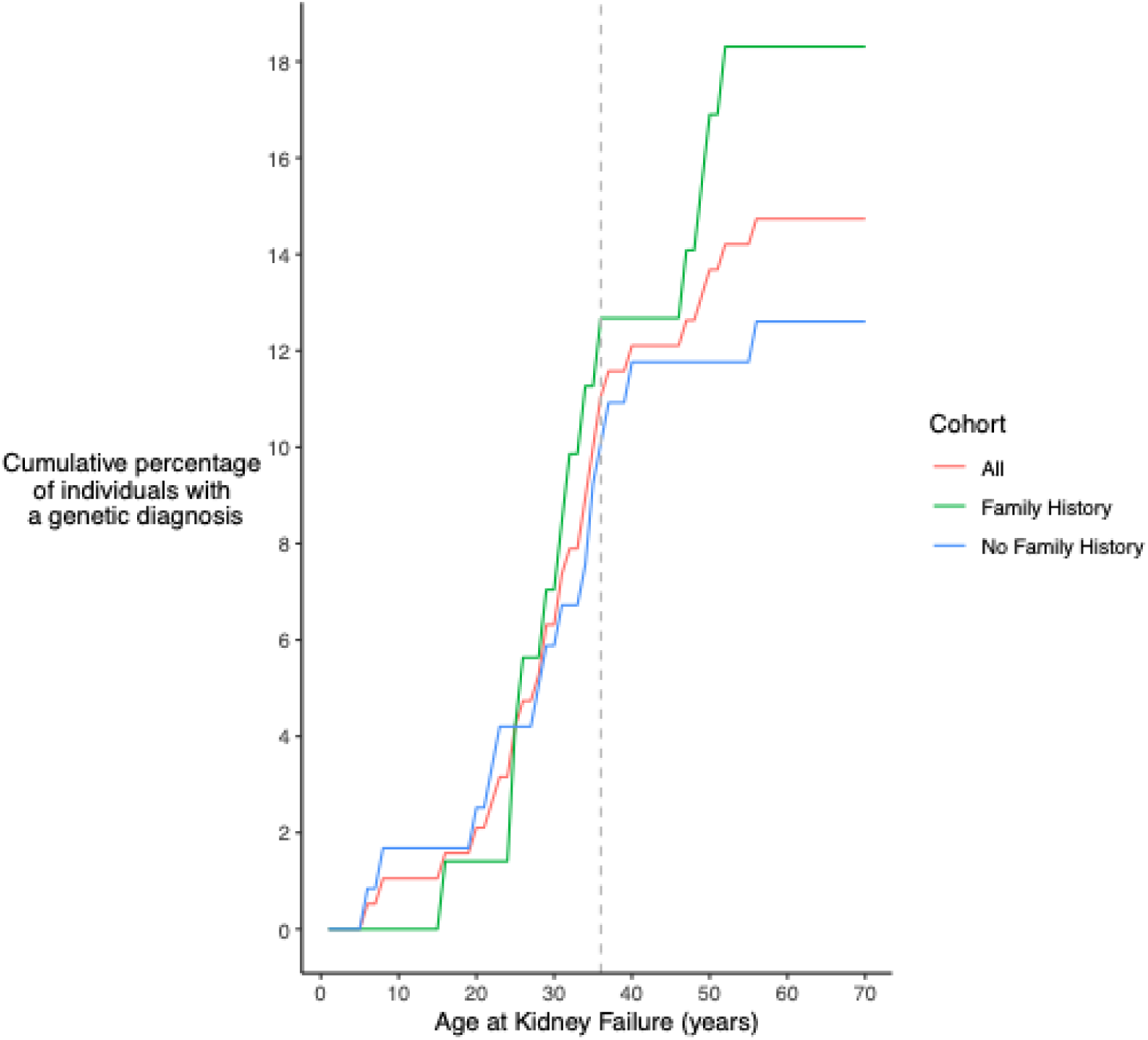
Proportion of patients with unexplained kidney failure with a genetic diagnosis stratified by family history. The grey dashed line represents the median age of kidney failure in the cohort of 36 years.

Three individuals with no family history who developed KF ≥36 years old received a genetic diagnosis. Interestingly, all three were heterozygous for type IV collagen variants: a likely pathogenic missense C*OL4A3*:c.3760G>C;p.(Gly1254Arg) variant, previously reported in a patient with Alport syndrome, was detected in a female with diabetes mellitus who developed KF in their late 30s (36-40); a pathogenic frameshift *COL4A4*:c.282_283del;p.(Asp96ProfsTer13) variant was identified in a male who KF in their 50s (50-55), who had microscopic haematuria, proteinuria, and gout; and a likely pathogenic *COL4A5*:c.367G>A;p.(Gly123Arg) variant was seen in a male with haematuria and proteinuria who reached KF in their 30s (36-40), consistent with a diagnosis of X-linked Alport syndrome. Although none of the individuals had a documented family history, identification of these type IV collagen variants initiated additional screening of family members.

We next looked across the exome for enrichment of rare variation in the unsolved uKF cohort to identify novel candidate genes. However, this collapsing gene-based analysis did not reveal any exome-wide significant genes at single nucleotide or structural variant level including collapsing variants that were in intronic, UTR or splicing domains (Supplementary Figure S3).

### High-risk *APOL1* genotype is associated with uKF in patients with recent African ancestry

High-risk *APOL1* genotypes are observed in 13% of individuals with recent African ancestry and have been associated with an increased risk of kidney disease, usually in an interferon-dependent manner^21–25^. We therefore hypothesized that these renal risk alleles might contribute to uKF in this cohort. Within the cohort, 27 probands were of African ancestry. 16 had biallelic *APOL1 G1/G2* risk alleles but two uKF patients carried the protective p.N264K variant alongside the G2 allele, leaving 14 cases with high-risk *APOL1* genotype. High-risk *APOL1* genotypes were significantly enriched in individuals with uKF (14/27; 52%) when compared to controls of African ancestry (51/608; 8.4%; OR=11.67; 95%CI 4.80-28.62; *P=2.50×10^-8^*). Three uKF cases with high-risk *APOL1* genotype had been given a monogenic diagnosis (*NPHP1*,*EVC,COL4A4*). There was no significant difference in the age at KF between the high-risk *APOL1* and low-risk *APOL1* uKF cases of African ancestry (median 37 vs 39 years; *P=0.73*).

### Polygenic risk scoring for CKD and glomerular disease does not enrich in the uKF cohort

We next explored whether individuals with uKF had a genetic susceptibility to other non-monogenic kidney diseases applying PRSs for IgA nephropathy, SSNS, membranous nephropathy and CKD. Using a multivariable logistic regression model for each PRS in the absence or presence of high-risk *APOL1* genotype (with the top ten principal components, age, and sex as covariates) revealed high-risk *APOL1* genotype was strongly associated with uKF (Figure 3;*P=7.99×10^-8^*; OR=9.15; 95% CI 4.11-2.03). Unsolved KF cases had a lower IgA nephropathy PRS than controls (OR=0.87; 95%CI 0.76-0.99; *P=0.04*). There was no difference between unsolved KF cases and controls when applying PRSs for SSNS (*P=0.05*), membranous nephropathy (*P=0.63*) or CKD (*P=0.59*).

**Figure 3.**
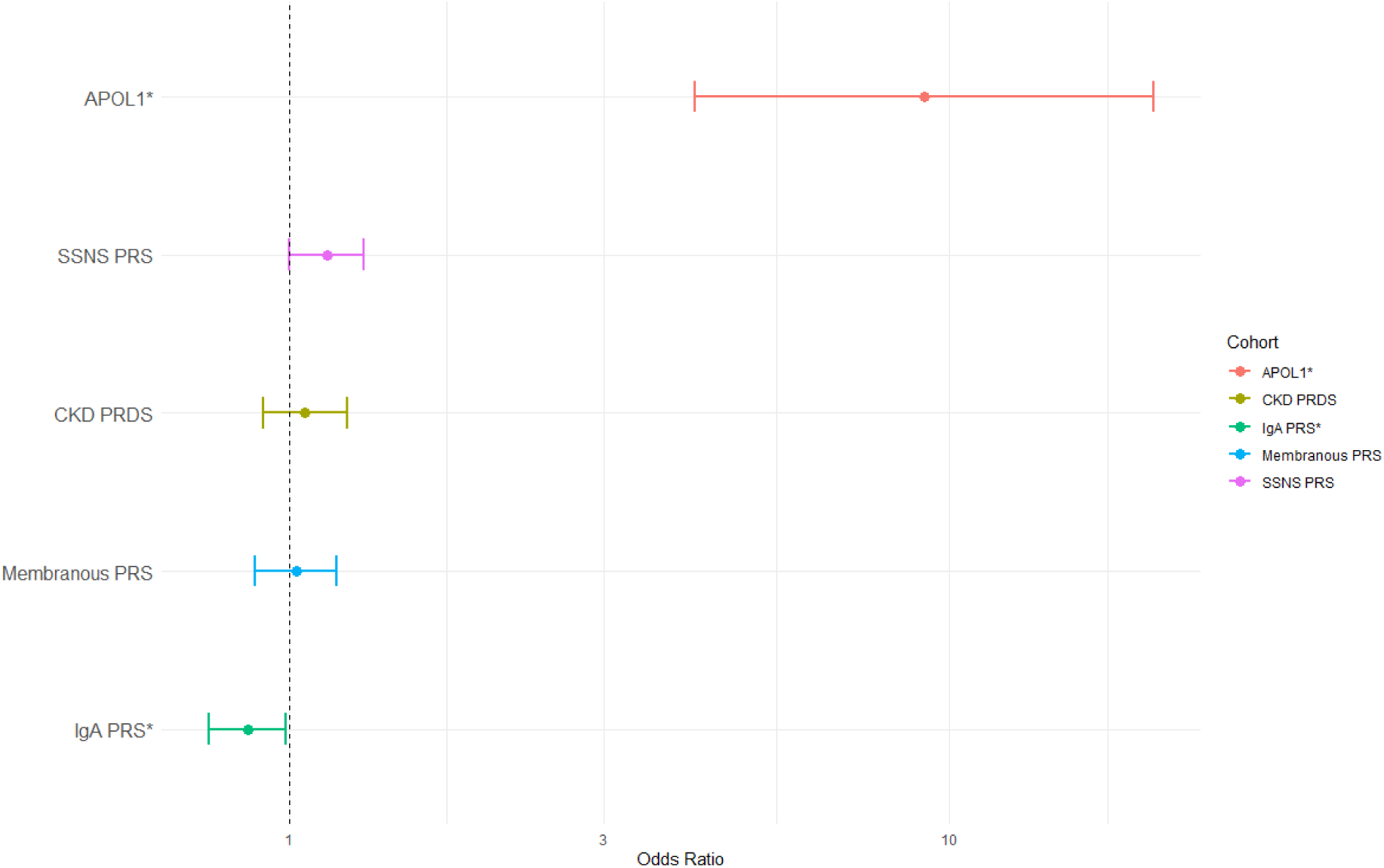
Odds Ratio of developing unexplained kidney failure across different polygenic risk scores and the presence of a high risk APOL1 genotype. * Denotes statistical significance.

### Individuals with the high-risk *APOL1* genotype have a higher SSNS PRS

Dividing the uKF cohort into those with a monogenic diagnosis (n=35), those with the high-risk *APOL1* genotype (n=14) and those with neither (unsolved uKF, n=169), the SSNS PRS was significantly higher in the high-risk *APOL1* genotype cohort (*P=6.16×10^-03^*) (Figure 4) when compared to the other case cohorts and controls. There was no difference in the remaining glomerular or CKD PRSs.

**Figure 4.**
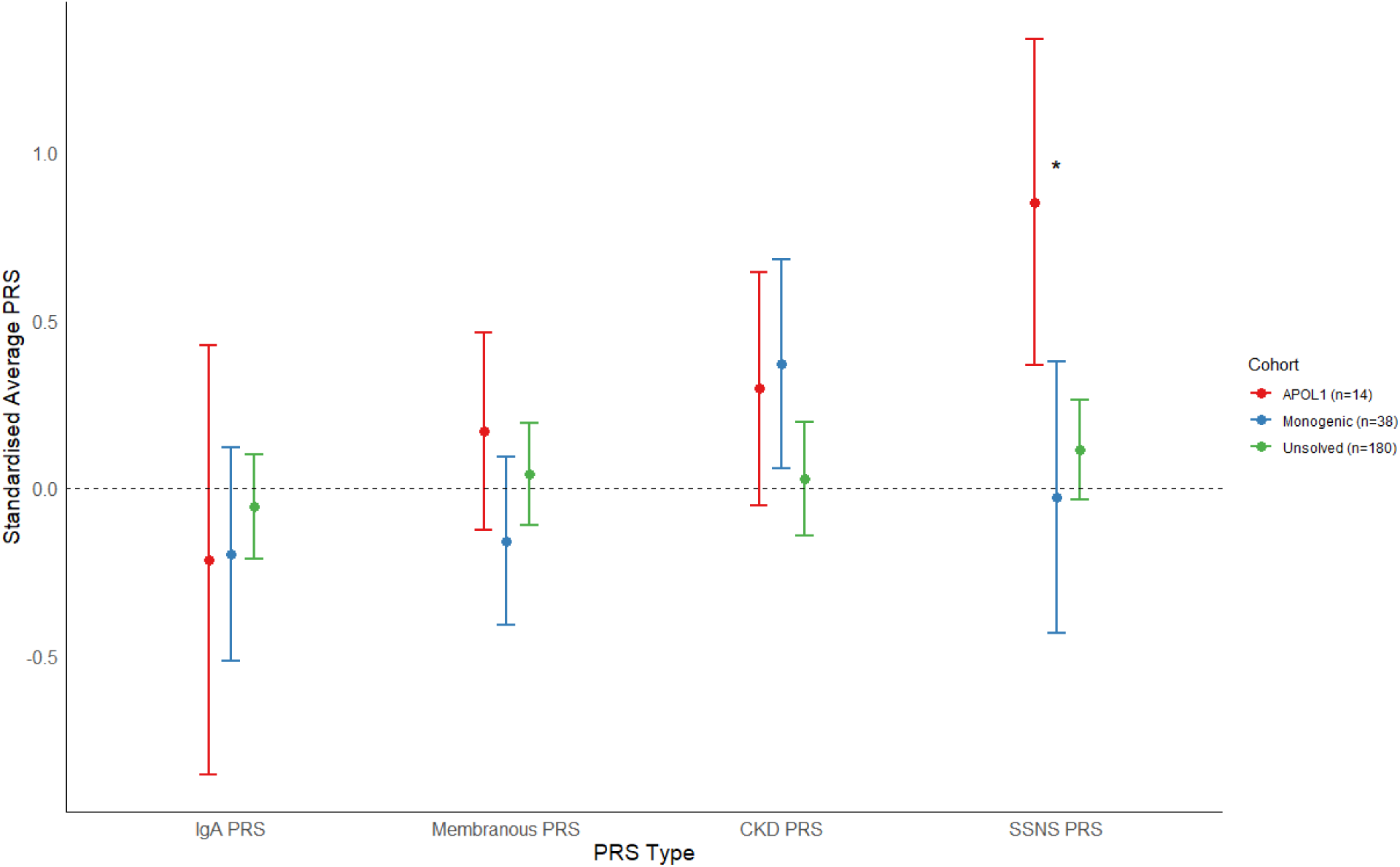
Average polygenic risk score by cohort and per PRS type. Each point is the average PRS with the bars representing the 95% confidence interval. Dotted line represents the normalized control PRS scores. * Denotes statistical significance. PRS – polygenic risk score, SSNS – steroid sensitive nephrotic syndrome, CKD – chronic kidney disease, IgA – Immunoglobulin A nephropathy

The SSNS PRS remained significantly elevated when comparing the high-risk *APOL1* genotype uKF cohort (n=14) and African ancestry controls with (P=0.048, n=51) and without (P=0.033, n=199) high-risk *APOL1* genotypes.

### Affected individuals with the high-risk *APOL1* genotype are enriched for *HLA-DQB1* variants found in West Africa

Three of the five loci contributing to the nephrotic syndrome PRS are in *HLA-DQB1*. To determine whether an HLA-risk allotype could be identified, we conducted a joint analysis of imputed HLA types across the UKBB and 100KGP in 21 individuals with early-onset KF and 468 controls, all with high-risk *APOL1* genotype. This revealed significant enrichment of HLA-DQB1*03:19 (P=0.001; OR=23.62; 95%CI 3.09-180.38, Supplementary Table 5).

This allele has a high frequency in Gambia (28%) with the next highest population frequency seen in Tanzania (5.1%)^26^. A full list of population allele frequencies can be found in Supplementary Table 5.

## Discussion

The monogenic diagnosis rate of 17% we observed in 218 patients with young-onset uKF is similar to the 17.1% reported using exome sequencing in the largest cohort investigated to date (n=281)^4^. Diagnostic rates of 12-47% have been reported in other exome-based studies focusing on patients with uKF and/or CKD^5,7,8^ and a recent WGS study in 100 individuals with CKD5 and median age of onset of 32 years reported a diagnostic yield of 25%^9^. Each study has differing proportions of familial disease and extra-renal features and used different gene panels, likely explaining some or all of this variability. WGS enables ascertainment of almost all types of genomic variation in both coding and non-coding regions and provides more uniform coverage across the genome offering enhanced detection of single-nucleotide variants, structural variants and those found in homologous pseudogenes, as in the case of *PKD1*. In the UK, WGS is currently available through the NHS Genomic Medicine Service for any individual with uKF under the age of 36 years, although high sequencing costs mean it is not routinely used in clinical practice in most health-care settings. Our results suggest prioritizing genetic testing for those with uKF occurring before the age of 36 years and/or a family history of kidney disease.

Beyond diagnostic yield, WGS also enables analysis of other types of genetic contributors to risk, including application of PRSs and modelling these alongside *APOL1* genotype and monogenic causes of KF. A PRS for CKD was not elevated in uKF cases, highlighting differences in genetic architecture in those who present with young-onset KF versus a general CKD population with progressive decline in GFR ^17,27^.

The glomerular disease PRS results should be interpreted within the context of how they were originally derived. For example, the IgA nephropathy PRS was developed in patients who are likely to have presented before CKD5 and the attenuated risk seen in uKF patients was mostly seen in those with a monogenic diagnosis or high-risk *APOL1* genotype (Figure 4). This suggests that undiagnosed IgA nephropathy is unlikely to explain a large proportion of this unsolved uKF cohort. Membranous nephropathy has a median age at diagnosis of 56.4 years and typically presents with nephrotic syndrome and preserved kidney function^2^. It is therefore unsurprising that there is no increased genetic risk for membranous nephropathy in this uKF cohort in which the upper age limit for KF was 50 years.

The *APOL1-*associated uKF patients did show enrichment for the SSNS PRS. Although derived from a genome-wide association study in childhood SSNS^28^, this PRS is similarly enriched in individuals with gene test negative steroid resistant nephrotic syndrome and adult-onset nephrotic syndrome and minimal change disease, suggesting that the score identifies genetic predisposition to autoimmune podocytopathy^14^. One possible explanation for this enrichment in individuals with *APOL1*-associated uKF is that the combination of *APOL1* high-risk genotype with autoimmune podocytopathy is a particularly strong risk factor for developing KF. However, none of the individuals included in the uKF cohort had a preceding diagnosis of nephrotic syndrome (or indeed proteinuric kidney disease) so data to support this hypothesis is lacking at present^29^. A potential role for autoimmune podocytopathy is further supported by our finding of a strong HLA association at DQB1 in those patients with KF and *APOL1* high-risk genotypes across both the UKBB and 100KGP cohorts. This HLA allele is seen at high frequency in West African populations and its enrichment in those with KF may result from population differences that were not adequately corrected by the genomic control measures we implemented. Interestingly, while previous studies have shown associations between SSNS and HLA-DQA1^29^ and DQB1^30^, this is the first time enrichment in these HLA alleles has been reported in individuals with a high-risk *APOL1* genotype and KF.

Our study is not without limitations. While we were able to study common and rare variants in a relatively large cohort of patients with uKF the number of individuals is too small to detect weaker genetic effects typically observed with PRS, especially when subgroup analyses were performed. Therefore, the lack of PRS enrichment for CKD, IgA and membranous nephropathy does not exclude the possibility that these causes of kidney disease are responsible for KF in a proportion of those presenting with uKF. In addition, we recognise that there are limitations to short-read WGS testing in both the detection of complex structural variants and the identification of variants in highly repetitive GC-rich regions, such as in the case of ADTKD-*MUC1,* and the use of long-read sequencing technologies in the future may improve the diagnostic yield in this patient group.

Our analyses confirm previous reports demonstrating a high proportion of monogenic uKF in younger people and reveal a strikingly high burden of high-risk *APOL1* genotypes among people with recent African ancestry in the UK with uKF. In addition, the enrichment of common genetic risk factors for SSNS in this latter group suggests that co-existence of more than one genetic risk or podocyte susceptibility factor in an individual may manifest clinically as uKF.

## Supporting information

supplementary methods and images

## Acknowledgments

This research was made possible through access to the data and findings generated by the 100,000 Genomes Project. The 100,000 Genomes Project is managed by Genomics England Limited (a wholly owned company of the Department of Health and Social Care). The 100,000 Genomes Project is funded by the National Institute for Health Research and NHS England. The Wellcome Trust, Cancer Research UK and the Medical Research Council have also funded research infrastructure. The 100,000 Genomes Project uses data provided by patients and collected by the National Health Service as part of their care and support. The authors gratefully acknowledge the participation of the patients, and their families recruited to the 100,000 Genomes Project.

## Author Contributions

DPG conceived the study. OSA, MMYC, GTD, KT conducted the analyses. OSA and MMYC wrote the manuscript with extensive input from DPG and amendments from GTD.

## Data Availability

All collapsing gene analyses results can be found in the Supplementary Tables (3-11). Details of the aggregated dataset used for the analysis can be found at: https://re-docs.genomicsengland.co.uk/aggv2/

The principal components were derived using the common, high quality independent SNPs derived by Genomics England: https://re-docs.genomicsengland.co.uk/principal_components/

Genomic and phenotype data from participants recruited to the 100,000 Genomes Project can be accessed by application to Genomics England Ltd (https://www.genomicsengland.co.uk/about-gecip/joining-research-community/).

The SSNS polygenic risk score can be found at: https://www.pgscatalog.org/score/PGS003354/

The membranous nephropathy polygenic risk score can be found at: https://www.ncbi.nlm.nih.gov/pmc/articles/PMC10116192/

The IgA polygenic risk score can be found at: https://www.columbiamedicine.org/divisions/kiryluk/resources.php

The CKD polygenic risk score can be found at: https://www.columbiamedicine.org/divisions/kiryluk/study_GPS_CKD.php

## Code Availability

Code used for the analyses in this paper can be found at: https://github.com/oalavijeh/unexplained_kf_paper.

Details of the rare variant workflow can be found at: https://re-docs.genomicsengland.co.uk/avt/

Details of the common variant GWAS workflow can be found at: https://re-docs.genomicsengland.co.uk/gwas/

## References

1. Kramer A, Pippias M, Noordzij M, et al. The European renal association - European dialysis and transplant association (ERA-EDTA) Registry Annual Report 2016: A summary. Clin Kidney J. 2019;12(5):702–720.

2. Wong K, Pitcher D, Braddon F, et al. Effects of rare kidney diseases on kidney failure: a longitudinal analysis of the UK National Registry of Rare Kidney Diseases (RaDaR) cohort. Lancet. Published online March 13, 2024. doi:10.1016/S0140-6736(23)02843-X

3. Connaughton DM, Hildebrandt F. Personalized medicine in chronic kidney disease by detection of monogenic mutations. Nephrol Dial Transplant. 2020;35(3):390–397.

4. Groopman EE, Marasa M, Cameron-Christie S, et al. Diagnostic Utility of Exome Sequencing for Kidney Disease. N Engl J Med. 2019;380(2):142–151.

5. Lata S, Marasa M, Li Y, et al. Whole-exome sequencing in adults with chronic kidney disease: A pilot study. Ann Intern Med. 2018;168(2):100–109.

6. Snoek R, van Jaarsveld RH, Nguyen TQ, et al. Genetics-first approach improves diagnostics of ESKD patients <50 years old. Nephrol Dial Transplant. 2022;37(2):349–357.

7. Connaughton DM, Kennedy C, Shril S, et al. Monogenic causes of chronic kidney disease in adults. Kidney Int. 2019;95(4):914–928.

8. Ottlewski I, Münch J, Wagner T, et al. Value of renal gene panel diagnostics in adults waiting for kidney transplantation due to undetermined end-stage renal disease. Kidney Int. 2019;96(1):222–230.

9. Mallawaarachchi AC, Fowles L, Wardrop L, et al. Genomic Testing in Patients with Kidney Failure of an Unknown Cause: a National Australian Study. Clin J Am Soc Nephrol. Published online May 3, 2024. doi:10.2215/CJN.0000000000000464

10. Richards S, Aziz N, Bale S, et al. Standards and guidelines for the interpretation of sequence variants: a joint consensus recommendation of the American College of Medical Genetics and Genomics and the Association for Molecular Pathology. Genet Med. 2015;17(5):405–424.

11. Chen X, Schulz-Trieglaff O, Shaw R, et al. Manta: rapid detection of structural variants and indels for germline and cancer sequencing applications. Bioinformatics. 2016;32(8):1220–1222.

12. Roller E, Ivakhno S, Lee S, Royce T, Bioinformatics ST., 2016 U. Canvas: versatile and scalable detection of copy number variants. academic.oup.com. https://academic.oup.com/bioinformatics/article-abstract/32/15/2375/1743834

13. Gupta Y, Friedman DJ, McNulty MT, et al. Strong protective effect of the APOL1 p.N264K variant against G2-associated focal segmental glomerulosclerosis and kidney disease. Nat Commun. 2023;14(1):7836.

14. Kiryluk K, Sanchez-Rodriguez E, Zhou XJ, et al. Genome-wide association analyses define pathogenic signalling pathways and prioritize drug targets for IgA nephropathy. Nat Genet. 2023;55(7):1091–1105.

15. Gupta S, Downie ML, Cheshire C, et al. A Genetic Risk Score Distinguishes Different Types of Autoantibody-Mediated Membranous Nephropathy. Glomerular Dis. 2023;3(1):116–125.

16. Downie ML, Gupta S, Chan MMY, et al. Shared genetic risk across different presentations of gene test-negative idiopathic nephrotic syndrome. Pediatr Nephrol. Published online November 10, 2022. doi:10.1007/s00467-022-05789-7

17. Khan A, Turchin MC, Patki A, et al. Genome-wide polygenic score to predict chronic kidney disease across ancestries. Nat Med. 2022;28(7):1412–1420.

18. Hinrichs AS, Karolchik D, Baertsch R, et al. The UCSC Genome Browser Database: update 2006. Nucleic Acids Res. 2006;34(Database issue). doi:10.1093/NAR/GKJ144

19. Chang CC, Chow CC, Tellier LCAM, Vattikuti S, Purcell SM, Lee JJ. Second-generation PLINK: Rising to the challenge of larger and richer datasets. Gigascience. 2015;4(1):7.

20. Wickham H. Ggplot2. Springer New York

21. Kopp JB, Nelson GW, Sampath K, et al. APOL1 genetic variants in focal segmental glomerulosclerosis and HIV-associated nephropathy. J Am Soc Nephrol. 2011;22(11):2129–2137.

22. Genovese G, Friedman DJ, Ross MD, et al. Association of trypanolytic ApoL1 variants with kidney disease in African Americans. Science. 2010;329(5993):841–845.

23. Friedman DJ, Kozlitina J, Genovese G, Jog P, Pollak MR. Population-based risk assessment of APOL1 on renal disease. J Am Soc Nephrol. 2011;22(11):2098–2105.

24. Foster MC, Coresh J, Fornage M, et al. APOL1 variants associate with increased risk of CKD among African Americans. J Am Soc Nephrol. 2013;24(9):1484–1491.

25. Kasembeli AN, Duarte R, Ramsay M, et al. APOL1 risk variants are strongly associated with HIV-associated nephropathy in black South Africans. J Am Soc Nephrol. 2015;26(11):2882–2890.

26. Gonzalez-Galarza FF, McCabe A, Santos EJMD, et al. Allele frequency net database (AFND) 2020 update: gold-standard data classification, open access genotype data and new query tools. Nucleic Acids Res. 2020;48(D1):D783–D788.

27. Vy HMT, Coca SG, Sawant A, et al. Genome-Wide Polygenic Risk Score for CKD in Individuals with APOL1 High-Risk Genotypes. Clin J Am Soc Nephrol. 2024;19(3):374–376.

28. Dufek-Kamperis S, Kleta R, Bockenhauer D, Gale D, Downie ML. Novel insights in the genetics of steroid-sensitive nephrotic syndrome in childhood. Pediatr Nephrol. 2021;36(8):2165–2175.

29. Adeyemo A, Esezobor C, Solarin A, et al. HLA-DQA1 and APOL1 as Risk Loci for Childhood-Onset Steroid-Sensitive and Steroid-Resistant Nephrotic Syndrome. Am J Kidney Dis. 2018;71(3):399–406.

30. Barry A, McNulty MT, Jia X, et al. Multi-population genome-wide association study implicates immune and non-immune factors in pediatric steroid-sensitive nephrotic syndrome. Nat Commun. 2023;14(1):1–13.

