## supplementary methods and images for "Quantifying the genetic contributions in unexplained kidney failure reveals *APOL1-HLA* interaction"

### Supplementary Methods 1: DNA preparation, and whole genome sequencing

#### DNA preparation and extraction

99% of DNA samples were extracted from blood and prepared using EDTA, with the remaining 1% sourced from saliva, tissue and fibroblasts. Libraries were prepared using the Illumina TruSeq DNA PCR-Free High Throughput Sample Preparation kit or the Illumina TruSeq Nano High Throughput Sample Preparation kit.

#### Whole-genome sequencing, alignment and variant calling

Samples were sequenced with 150bp paired-end reads using an Illumina HiSeq X instrument and processed on the Illumina North Star Version 4 Whole Genome Sequencing Workflow (NSV4, version 2.6.53.23), comprising the iSAAC Aligner (version 03.16.02.19) and Starling Small Variant Caller (version 2.4.7). Samples were aligned to the Homo Sapiens NCBI GRCh38 assembly. Samples achieved a mean of 97.38% coverage at 15X with a median genome-wide coverage of 39.11X. Samples with <2% cross-contamination as determined by the VerifyBamID algorithm were kept. Copy number and structural variant (>50bp) calling was performed using CANVAS^1^ (version 1.3.1) and MANTA^2^ (version 0.28.0) respectively. CANVAS determines coverage and minor allele frequencies to assign copy number (>10kb) whereas MANTA combines paired and split-read algorithms to detect structural variants (< 10kb).

### Supplementary Methods 2: Genomic variant calling format file annotation and variant-level quality control

All genomic files were aligned to the GRCh38 reference human genome. Genomic variant call format files (gVCFs) were aggregated using gvcfgenotyper (Illumina, version: 2019.02.26) with variants normalized and multi-allelic variants decomposed using vt (version 0.57721). Variants were retained if they passed the following filters: missingness ≤ 5%, median depth ≥ 10, median GQ ≥ 15, percentage of heterozygous calls not showing significant allele imbalance for reads supporting the reference and alternate alleles (ABratio) ≥ 25%, percentage of complete sites (completeGTRatio) ≥ 50% and P value for deviations from Hardy-Weinberg equilibrium (HWE) in unrelated samples of inferred European ancestry ≥ 1×10-5. Male and female subsets were analysed separately for sex chromosome quality control. Per-variant minor allele count (MAC) was calculated across the case-control cohort, MAC is defined as the number of minor alleles counted for each marker. Annotation was performed using Variant Effect Predictor^3^ (VEP, version 98.2) including CADD^4^ (version 1.5), and allele frequencies from publicly available databases including gnomAD^5^ (version 3) and TOPMed^6^ (Freeze 5). Variants were filtered using bcftools^7^ (version 1.11).

### Supplementary Methods 3: Creation of ancestry matched unsolved cohort

#### Relatedness estimation and principal components analysis

All unsolved uKF cases were matched by genetically defined ancestry to a control cohort of 27,660 unaffected relatives of non-renal rare disease participants, excluding those with human phenotype ontology (HPO)^8^ terms and/or hospital episode statistics (HES) data consistent with kidney disease or failure. Given the small number of cases, we chose to analyse individuals from diverse backgrounds together, thereby preserving sample size and boosting power. To mitigate confounding due to population differences whilst using this mixed ancestry approach we employed two strategies, as previously described^9,10^. First, we carried out ancestry-matching of cases and controls using weighted principal components and second, utilized a generalized logistic mixed model to account for relatedness between individuals (detailed in the rare variant collapsing analysis section below).

We generated the principle components using a set of 127,747 high quality autosomal LD-pruned biallelic single nucleotide variants (SNVs) with a minor allele frequency (MAF) > 1% using PLINK^11^ (v1.9), MAF is defined as the frequency at which the second most common allele occurs in a given population. SNVs were included if they met all the following criteria: missingness < 1%, median GQ ≥ 30, median depth ≥ 30, AB Ratio ≥ 0.9, completeness ≥ 0.9. Ambiguous SNVs (AC or GT) and those in a region of long-range high LD (Linkage Disequilibrium) were excluded. LD pruning was carried out using an r2 threshold of 0.1 and window of 500kb. SNVs out of HWE in any of the AFR, EAS, EUR or SAS 1000 Genomes populations were removed (pHWE < 1 ×10-5). Using this variant set, a pairwise kinship matrix was generated using the PLINK2 implementation of the KING-Robust algorithm^12^ and a subset of unrelated samples was ascertained using a kinship coefficient threshold of 0.0884 (2nd degree relationships). Ten principal components were generated using PLINK^11^ for ancestry-matching and as covariates in the association analyses.

#### Ancestry matching

Given the mixed-ancestry composition of the cohort we employed a case-control ancestry-matching algorithm which uses the generated principle components to optimize genomic similarity and minimize the effects of population structure as previously described^9,10,13^ with each case having to match a minimum of two controls to be included in the final cohort. The final cohort consisted of187 unsolved uKF patients and 26,373 ancestry matched controls (Supplementary Figure 1).

After quality control, relatedness filtering and ancestry we had a cohort of 187 unsolved uKF patients and 26,373 ancestry-matched controls.

### Supplementary Methods 4: Rare variant selection for collapsing analysis.

Single variant association testing is underpowered when variants are rare and a collapsing approach which aggregates variants by gene or genomic region can be adopted to boost power. Whilst the variants are aggregated by gene, they are also filtered to help boost power. Each set of filters are applied as a “mask” which instructs the workflow to collapse variants as per a set of parameters. The mask used for this analysis were a rare, damaging missense mark (“missense+”), a high confidence loss of function mask (“LoF”), an intronic mask (“intronic”), a splice site mask, a 3-prime untranslated region mask (3’-UTR) and a 5-primte UTR mask (5’-UTR).

For the missense+ mask we extracted coding SNVs and indels with MAF < 0.01% in gnomAD annotated with one of the following: missense, in-frame insertion, in-frame deletion, start loss, stop gain, frameshift, splice donor, splice acceptor for each gene and further filtered them by CADD^4^ (v1.5) score using a threshold of ≥20 corresponding to the top 1% of all predicted deleterious variants in the genome. Variants meeting the following quality control filters were retained: MAC ≤ 20, median site-wide sequencing depth in non-missing samples > 20 and median GQ ≥ 30. Sample-level QC metrics for each site were set to minimum depth per sample of 10, minimum GQ per sample of 20 and ABratio P value > 0.001.

Variants with significantly different missingness between cases and controls (P<10-5) or >5% missingness overall were excluded. For the dedicated loss-of-function (LoF) analysis, variants were selected that were deemed to be of “high confidence” (HC) by the loss-of-function transcript effect estimator (LOFTEE)^5^. These assessed variants are stop-gained, splice site disrupting or frameshifts and collapsed on a per-gene basis. The same quality controls were applied as above. For the splice site mask, variants were collapsed if their SpliceAI^14^ scores were greater than 0.8 (on a scale of 0-1) indicating variants that were highly likely to affect splicing. These were divided by their predicted effect on splicing (donor loss, donor gain, acceptor loss and acceptor gain). The intronic mask was defined by variants labelled as “intronic” by VEP with a CADD score greater than 20 and with a MAF<0.01% in gnomAD. The 5’ and 3’ UTR masks were defined by variants with a MAF<0.01% in gnomAD, a CADD score greater than 10 and found within their respective UTR region as labelled by VEP.

All masks were applied to the uKF cohort and controls. Association testing was performed using the Scalable and Accurate Implementation of Generalized mixed model (SAIGE-GENE) (v0.42.1)^15^ to ascertain whether rare variation was enriched in cases on a per-gene basis exome-wide. SAIGE-GENE uses a generalized mixed-model to correct for population stratification and cryptic relatedness as well as a saddle point approximation and efficient resampling adjustment to account for the inflated type 1 error rates seen with unbalanced case-control ratios. It combines single-variant score statistics and their covariance estimate to perform SKAT-O^16^ gene-based association testing, upweighting rarer variants using the beta (1,25) weights option. SKAT-O is a combination of a traditional burden and variance-component test and provides robust power when the underlying genetic architecture is unknown.

Sex, age and the top ten principal components were included as fixed effects when fitting the null model.

A Bonferroni adjusted P-value of 2.58×10^-6^ (0.05/19,364 genes) was used to determine the exome-wide significance threshold.

### Supplementary Methods 5: Structural and copy number variant analysis.

Structural variants (SV) were called from WGS using the Genomics England pipeline that incorporates CANVAS^1^ to detect copy number (>10kb) and MANTA^17^ to identify SVs greater than 50bp. CANVAS uses read depth to assign copy number variant (CNV) losses and gain. MANTA uses both discordant read-pair and split-read data to identify SV regions.

The following quality control filters were applied to the variants: CNV length > 10kb and Q-score ≥ Q10 indicating 90% confidence there is a variant present, a quality score ≥ 20 indicating 99% confidence that there is a variant at the site, GQ ≥ 15 indicating 95% confidence that the genotype assigned to a sample is correct, and MaxMQ0Frac < 0.4 which indicates the proportion of uniquely mapped reads around either breakend. Variants without paired read support, inconsistent ploidy, or depth > 3x the mean chromosome depth near one or both breakends were excluded.

For each sample BEDTools^18^ was used to extract SVs that intersected at least one exon by a minimum of 1 base pair. Variants were then separated into CNV, deletion (DEL), duplication (DUP), and inversion (INV) sets before being filtered to remove common SVs of the same type.

SVs were removed if they had a minimum 70% reciprocal overlap with SVs^19^ with allele frequency >1% in gnomAD and/or a dataset of common (AF>0.1%) SVs generated from 12,243 cancer patients recruited to 100KGP. SVs were then merged using SURVIVOR^20^ allowing a maximum distance of 300bp between pairwise breakpoints and allele frequencies calculated using BCFtools^7^.

After removal of overlapping common variants, a custom Perl script (Dr Helen Griffin, Newcastle University) was used to calculate allele frequencies for each type of SV across the combined case-control cohort using bins of 10kb across the entire genome. SVs with an AF < 0.1% were retained for further analysis.

Exome-wide gene-based burden testing was carried out using custom R scripts stratified by SV type. SVs were aggregated across 19,005 autosomal protein-coding genes. A two-sided Fisher’s exact test was used to compare SVs in cases and controls under a dominant inheritance model. The Bonferroni correction for the number of genes (P=0.05/19,005=2.6x10-6) tested was applied, although with the knowledge that this is likely to be too stringent given the tests are not truly independent (one SV can affect multiple genes).

While MANTA can detect deletions and tandem duplications <10kb, inversions, and interchromosomal translocations, it cannot reliably identify dispersed duplications, small inversions (<200bp), fully assembled large insertions (>2x150bp) or breakends where repeat lengths approach the read size (150bp) . Very few insertions were identified in this cohort and in view of this they were excluded from downstream analysis. In addition, variants classified as translocations, single breakends or complex SVs which are more difficult to accurately resolve were filtered out.

### Supplementary Methods 6: HLA analysis in 100KGP and UKBB

Using HIBAG^21^, HLA types were imputed at two field resolution for HLA-A, HLA-C, HLA-B, HLA-DRB1, HLA-DQA1, HLA-DQB1, and HLA-DPB1 centrally by Genomics England^22^. These were then extracted for the cohort of patients with high risk APOL1 variants.

To increase numbers to detect whether HLA allotype modified APOL1 related risk, we sought additional cases and controls from the UK Biobank (UKBB). Using whole exome sequencing data in UKBB we identified individuals carrying high-risk APOL1 variants (G1/G1, G1/G2 and G2/G2) were identified. Cases (n=5) were defined as people with CKD4, CKD5 or requiring renal replacement therapy before the age of 50, based on hospital inpatient diagnoses and operative and procedural records. Controls (n=417) were people with the same age-range as cases without any hypertension, chronic kidney disease related codes. Imputed HLA allotypes (previously performed by the UK Biobank using HLA*IMP:02^23^) were extracted for all individuals and the top two alleles of each HLA type were assigned based on the highest imputation probabilities.

To address confounding by population stratification, we generated principal components of the common variant genotype matrix of the cohort, to include as covariates. A list of high quality, biallelic, LD- and complex-region-pruned SNPS with a minor allele frequency > 0.05, defined in the Genomics England dataset (see data availability) for ancestry estimation, were extracted. These were intersected with UK Biobank imputed genotypes (imputed genotypes provided by UK Biobank, previously imputed against TopMED R2 panel). PCA was performed on 63,523 SNPs using PLINK (version 1.9)^11^.

The HLA types in cases and controls with high risk APOL1 variants were then joint-analysed using a logistic regression model as implemented within the HIBAG tool using HLA-type and 5 principal components, a chi-squared test was used to attribute for significance using a Bonferroni corrected P-value of 0.007 (7 HLA classes analysed, α=0.05).

### Supplementary Figures

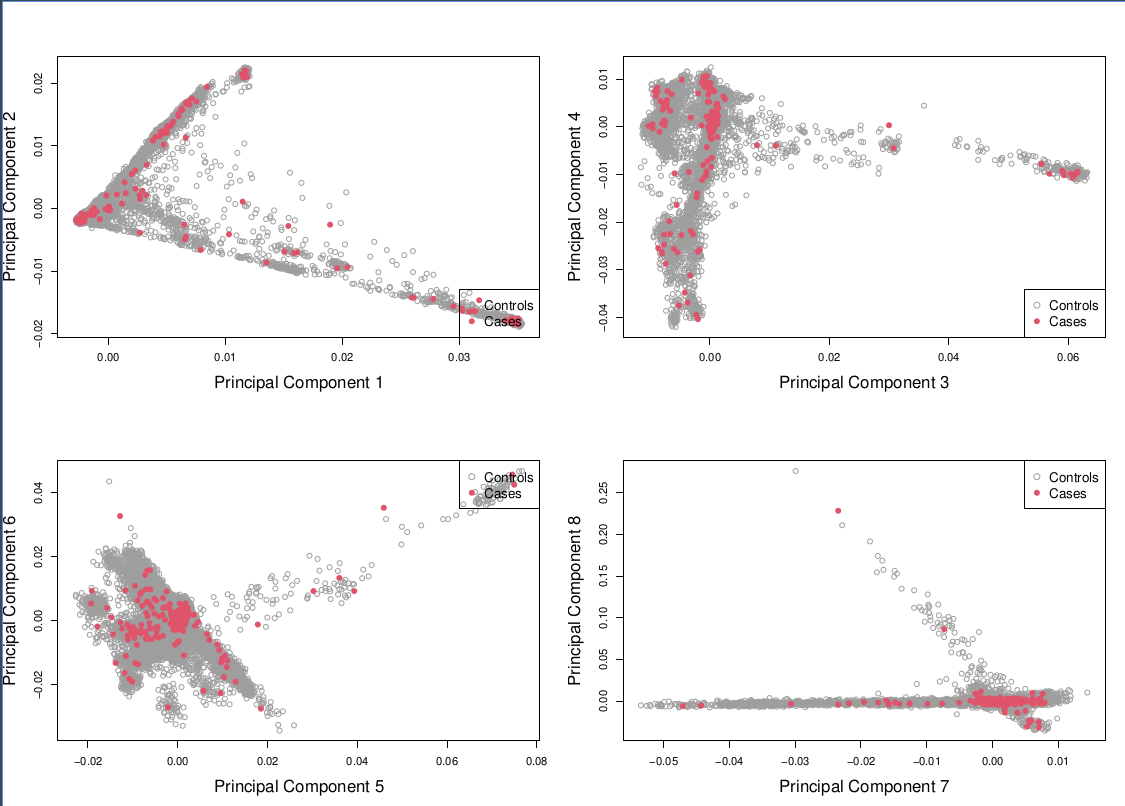

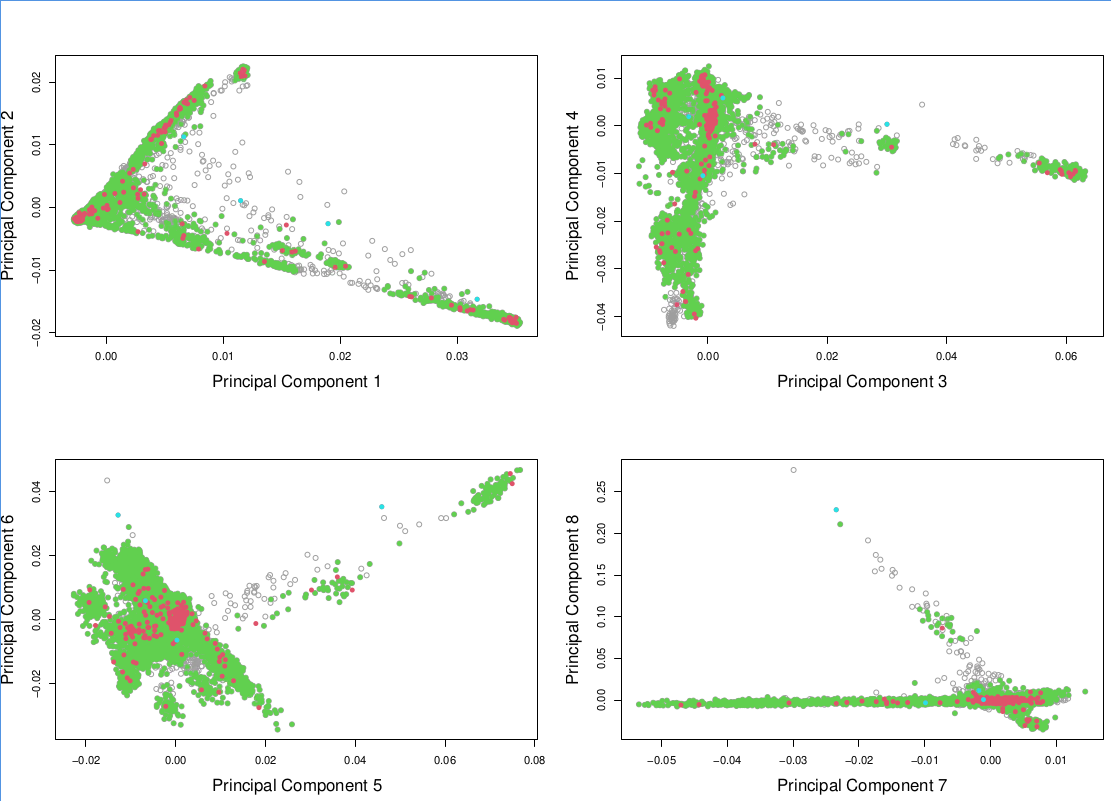

Supplementary Figure S1 – Ancestry Matching

1a. Principal component analysis showing the first eight principal components for matched cases (red) and controls (white). 1b. Principal component analysis showing the first eight principal components for matched cases (red) and controls (green) and unmatched cases(blue) and controls (grey). This highlights that cases are taken from multiple different ancestries with the appropriate matched controls.

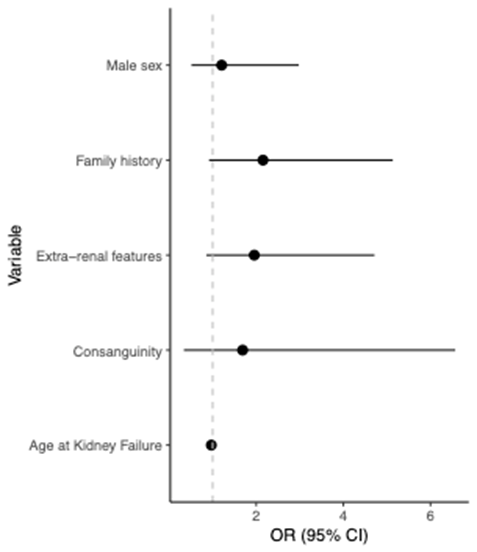

Supplementary Figure S2 - Multivariable logistic regression of factors associated with a positive genetic diagnosis. KF, kidney failure; OR, odds ratio; CI, 95% confidence interval.

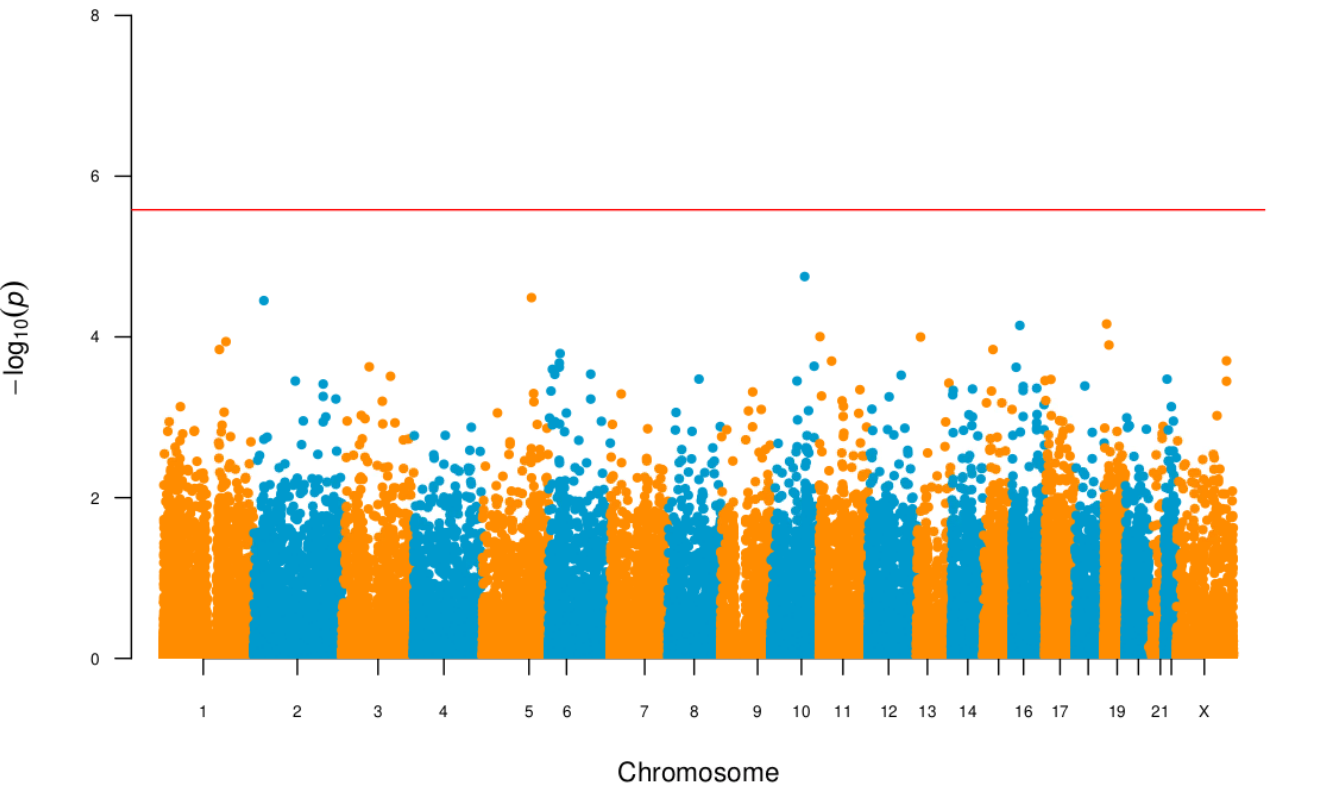

Supplementary Figure S3 – Aggregated Manhattan Plot of multiple collapsing gene-based tests

Gene based Manhattan. Each point is a gene made up of variants meeting the various collapsing masks described in Supplementary Methods 4 – Missense+, LoF, Splice side, intronic, 5’ and 3’-UTR as well as the exome-wide structural variants. The red line represents the genome wide significance line.

### References

1.	Roller, E. *et al.* Canvas: versatile and scalable detection of copy number variants. *academic.oup.com*.

2.	Chen, X. *et al.* Manta: rapid detection of structural variants and indels for germline and cancer sequencing applications. *Bioinformatics* **32**, 1220–1222 (2016).

3.	McLaren, W. *et al.* The Ensembl Variant Effect Predictor. *Genome Biol.* **17**, 1–14 (2016).

4.	Rentzsch, P., Witten, D., Cooper, G. M., Shendure, J. & Kircher, M. CADD: predicting the deleteriousness of variants throughout the human genome. *Nucleic Acids Res.* **47**, D886–D894 (2019).

5.	Karczewski, K. J. *et al.* The mutational constraint spectrum quantified from variation in 141,456 humans. *Nature* **581**, 434–443 (2020).

6.	Taliun, D. *et al.* Sequencing of 53,831 diverse genomes from the NHLBI TOPMed Program. *Nature* **590**, 290–299 (2021).

7.	Danecek, P. *et al.* Twelve years of SAMtools and BCFtools. *Gigascience* **10**, (2021).

8.	Köhler, S., Gargano, M., Research, N. M.-. …. A. & 2021, U. The human phenotype ontology in 2021. *academic.oup.com*.

9.	Sadeghi-Alavijeh, O. *et al.* Rare variants in the sodium-dependent phosphate transporter gene SLC34A3 explain missing heritability of urinary stone disease. *Kidney Int.* (2023) doi:10.1016/j.kint.2023.06.019.

10.	Chan, M. M. Y. *et al.* Diverse ancestry whole-genome sequencing association study identifies TBX5 and PTK7 as susceptibility genes for posterior urethral valves. *Elife* **11**, (2022).

11.	Purcell, S. *et al.* PLINK: a tool set for whole-genome association and population-based linkage analyses. *Am. J. Hum. Genet.* **81**, 559–575 (2007).

12.	Manichaikul, A. *et al.* Robust relationship inference in genome-wide association studies. *Bioinformatics* **26**, 2867–2873 (2010).

13.	Sadeghi-Alavijeh, O. *et al.* Quantifying variant contributions in cystic kidney disease using national-scale whole genome sequencing. *J. Clin. Invest.* (2024) doi:10.1172/JCI181467.

14.	Jaganathan, K. *et al.* Predicting Splicing from Primary Sequence with Deep Learning. *Cell* **176**, 535-548.e24 (2019).

15.	Zhou, W. *et al.* SAIGE-GENE+ improves the efficiency and accuracy of set-based rare variant association tests. *Nat. Genet.* **54**, 1466–1469 (2022).

16.	Lee, S. *et al.* Optimal unified approach for rare-variant association testing with application to small-sample case-control whole-exome sequencing studies. *Am. J. Hum. Genet.* **91**, 224–237 (2012).

17.	Chen, X. *et al.* Manta: rapid detection of structural variants and indels for germline and cancer sequencing applications. *Bioinformatics* **32**, 1220–1222 (2016).

18.	Quinlan, A. R. & Hall, I. M. BEDTools: a flexible suite of utilities for comparing genomic features. *Bioinformatics* **26**, 841–842 (2010).

19.	Collins, R. L. *et al.* A structural variation reference for medical and population genetics. *Nature* **581**, 444–451 (2020).

20.	Jeffares, D. C. *et al.* Transient structural variations have strong effects on quantitative traits and reproductive isolation in fission yeast. *Nat. Commun.* **8**, 14061 (2017).

21.	Zheng, X. *et al.* HIBAG--HLA genotype imputation with attribute bagging. *Pharmacogenomics J.* **14**, 192–200 (2014).

22.	Kousathanas, A. *et al.* Whole-genome sequencing reveals host factors underlying critical COVID-19. *Nature* **607**, 97–103 (2022).

23.	Dilthey, A. *et al.* Multi-population classical HLA type imputation. *PLoS Comput. Biol.* **9**, e1002877 (2013).
